# Understanding the impacts of reductions in local government expenditure on food safety services, England 2009/10 - 2019/20

**DOI:** 10.1101/2025.04.04.25325088

**Authors:** Lauren Murrell, Helen. E Clough, Roger Gibb, Xingna Zhang, Marie Chattaway, Mark. A Green, Iain Buchan, Benjamin Barr, Daniel Hungerford

## Abstract

**Background:** Environmental and Regulatory (ER) services, provided by local authorities, play a key role in public health protection. Food safety teams within ER services carry out regular food hygiene inspections, advice and education visits, and investigate foodborne disease outbreaks. Following the 2008 financial crisis, local authorities have faced substantial funding cuts. This study investigates the impact of local government expenditure reductions on food safety services.

**Methods:** We used fixed effects panel regression to analyse the effect of food safety expenditure reductions on food hygiene staffing levels, food hygiene interventions (e.g. inspection, sampling, advice and education), and the proportion of establishments rated food hygiene compliant. We also examined the effect of staff reductions on the number of interventions achieved and establishment compliance.

**Results:** A £1 decrease in food safety expenditure per capita was associated with a 2% (95% CI 0.7% to 3.3%) decrease in staffing levels and a 1.7% (95% CI 0.2% to 3.3%) decrease in the number of interventions achieved per establishment. One-unit reduction in staff was associated with a 49.2% (95% CI 18.7%, 87.6%) decrease in the number interventions achieved. We found no evidence of an association between expenditure or staff levels and the proportion of compliant establishments.

**Conclusion:** Spending reductions negatively impact the capacity of food safety teams to provide key services. Reductions in food safety expenditure significantly impact food hygiene staff levels and service provision. This raises concerns about the capacity of food safety teams to operate and the potential for increased public risk of gastrointestinal infections.

## Introduction

### Background

There are approximately 17 million cases of gastrointestinal infections annually in the UK (1), making them a significant public health concern. These illnesses often result in diarrhoea and vomiting, and in some cases death (2). Gastrointestinal infections are usually spread through contaminated food, water, and surfaces (3). There are an estimated 2.4 million cases of foodborne illnesses (4), resulting in 16,400 hospitalisations (2), and 180 deaths annually (5). Additionally, the total societal cost associated with foodborne disease is approximately £9.1 billion a year, including lost earnings, disturbance to business, medical costs, and costs associated with missed school (4).

In England, local authorities (LA) are responsible for enforcing food safety (6) and preventing foodborne illness and outbreak. Food safety is provided by environmental and regulatory (ER) services and is a statutory service, meaning it is a local authority’s duty to provide it. The Food Standards Agency (FSA) is the central authority that oversees food safety controls enforced by local authorities (6). These controls are delivered by Environmental Health (EH) staff (7), also called environmental health officers (EHO’s) or practitioners (EHP’s). Food safety services include food hygiene inspections to ensure food is safe from bacterial contamination (7), testing food samples, food safety advice, and investigation of food poisoning outbreaks and foodborne illness (8). The functions provided by ER services under food safety are likely important for prevention of gastrointestinal infections.

The financial crisis of 2008 saw austerity measures implemented by central government to control national debt. This resulted in funding cuts to local authority services across England with reductions in spending power of up to 50% (9). Although ER services are largely statutory, there have been substantial cuts to these services which disproportionately impacted poorer areas. Per capita spending on food safety and infection control services declined by 22.8% in the most deprived areas compared to 6.3% in the least deprived areas between 2009/10 and 2020/21 (10).

There are signs that local funding cuts are negatively impacting local authority services. However, many services lack data on outputs or outcomes, making it difficult to measure the impact of expenditure reductions (11). Since 2010 local authority services have seen staff reductions, including in food safety services (7,12). There has also been reduction in waste collection, library service points, bus services (11), reports of food standards, health and safety focusing on high-risk businesses, and scaling back proactive work that may prevent serious risks (13).

The extent to which local funding cuts have impacted food safety services is unreported. We aim to measure the impact of reductions in food safety expenditure on food hygiene staff, the number of interventions achieved and the proportion of broadly compliant establishments.

## Methods

We carried out a longitudinal study at local authority level using data for the financial years of 2009/10 to 2019/20. The study focused on England and used data from 314 lower tier local authorities, that are responsible for environmental services in England. In total 26 LAs were removed from final analysis. Thirteen LAs were excluded from the study due to missing food safety expenditure data. Five were excluded due to food hygiene returns being reported with food standards and a further six were removed due to issues with consistency in LA reporting. Ilses of Scilly and City of London were excluded due to low population sizes and distinct funding structure. The local authority geographical boundaries used in this study are 2020 boundaries. In instances where local authority data were not at this level, data were aggregated and transformed to 2020 level to provide consistent data for this period.

### Local authority data

### Expenditure data

Expenditure data were sourced from the Place-based Longitudinal Data Resource (PLDR) (14), which collates publicly available annual local authority outturn data for cultural, environmental and regulatory services reported to the UK central government’s department of housing and local government (15). We analyzed annual food safety expenditure, by financial year, defined as spending reported in budget outturns for environmental and regulatory services under the title of food safety. This was adjusted for inflation using gross domestic product deflator (16), then normalized by ONS mid-year population estimates to give food safety services expenditure per capita.

### Service Indicator data

We defined three local authority food safety service indicators. Firstly, the annual number of interventions carried out per establishment. Interventions include inspections, sampling, audits, monitoring, surveillance and intelligence gathering, as well as advice and education (17). Local authorities use a risk-based approach to inform intervention frequency and type, described in table 1. Data were obtained from the FSA’s local authority enforcement monitoring system (LAEMS) (18) through a freedom of information request. More information can be found in section 1 of the supplementary material.

**Table 1:** Further background information on data.

|  |  |  |  |
| --- | --- | --- | --- |
| **Food hygiene interventions** include inspections, sampling, audits, monitoring, surveillance and intelligence gathering as well as advice and education. Interventions are carried out by **Environmental Health staff**. |  |  |  |
| **The type and frequency of interventions is determined by the risk the establishment faces to the public** | **Risk rating** | **Frequency and intervention type** |  |
|  | A | </6 months | Inspection, partial inspection, or audit |
|  | B | </12 months | Inspection, partial inspection, or audit |
|  | C | </18 months | Inspection, partial inspection or audit (If broadly compliant interventions can alternate between the above and another type of official control |
|  | D | </24 months | Alternate between inspection, partial inspection or audit and other types of intervention |
|  | E | Every 3 years or alternative programme | Alternative enforcement strategy |
| **Food Hygiene rating score is a rating from 0-5 which reflects the standards of hygiene on the day of inspection. Only establishments that directly supply the public will receive a food hygiene rating score.**<br>**The food hygiene score is determined after inspection, audit or partial audit** |  |  |  |

Secondly, we sourced food hygiene full-time-equivalent (FTE) position figures from LAEMS, as a measure of staff per capita, normalized using ONS population estimates. This was to indicate resource level. The term staff or staffing levels will be used when referring to FTE. See supplementary material section 1.3 for more detail.

Thirdly, we defined the proportion of broadly compliant food establishments (cafes, restaurants, pubs, and food shops) (19) as the number of businesses that received a rating of three or greater, divided by the number of establishments rated that year. Food hygiene scores range from 0 to 5, reflecting the food hygiene standards found on the day of inspection (19). A score of 0 means urgent improvement is required, a score of 5 means the hygiene standards are very good (20) see section 1.4 of the supplementary material for more detail. We acquired this data from the Consumer Data Research Centre (CDRC) which sources and compiles data from the FSA (20). We aggregated data from business level to local authority level and transformed it to financial year for consistent reporting across all data. The FHRS data were analyzed for years 2012/13 to 2019/20 due to the implementation FHRS scheme in 2010 likely leading to reduced data for years 2010-2012.

### Missing data

There was missing data for Food Safety expenditure, with 58 local authorities having at least one year with no expenditure data reported. This could be due to reporting under another spending line, or local authorities not disaggregating at the required level. This may have also contributed to missing data in outcome variables of interest such as staff and interventions. In some instances, local authorities reported food standards returns only, or food standards combined with food hygiene returns; such local authorities were removed. We attributed local authorities missing data for FHRS to the roll out of the scheme in late 2010, as there was no missing data after 2012. Local authorities with more than 50% data missing were removed and multiple imputation was used to estimate the values for remaining missing data, see supplementary material (section 2) for more information.

### Analysis

Data were analysed descriptively by year, deprivation level, local authority type and rural or urban classification (see supplementary material section 3). We then used fixed effects panel regression models to estimate the change in number FTE per capita, number of interventions achieved per establishment, and the proportion of compliant establishments associated with each £1 decrease in food safety expenditure per capita. All models included a fixed effect for each local authority to remove all between local authority differences, so the models only estimate the association between exposure and outcomes within local authorities over time. All models also use robust clustered standard errors; this accounts for clustering within local authorities. All models included year as a continuous variable allowing us to control for temporal variation.

First, we modelled the number of FTE as the outcome, with food safety per capita as the exposure. We used a log linear regression model with the log population estimate as an offset to model FTE per capita. The model was weighted by the local authority population size. Secondly, we analysed the number of interventions per establishment as the outcome with food safety expenditure as the exposure. We used a log linear regression model with the number of establishments as the offset, to model the number of interventions per establishment. In addition, the model was weighted by the number of establishments in each local authority. We then modelled the number of interventions per establishment as the outcome with food safety per capita and FTE per 10,000 as the exposures. We used a log linear regression model with the log of the number of establishments as the offset. This model was weighted by the number of establishments.

We used binomial regression with logit link function to model the effect of food safety expenditure per capita on the proportion of broadly compliant establishments. This model was weighted by the total number of establishments rated. We repeated this model using the number of FTE per 10,000 as the main exposure, with the proportion of broadly compliant establishments as the outcome. Finally, we modelled FTE per 10,000 of the population and food safety expenditure per capita as the exposures on the proportion of compliant establishments.

## Results

Table 2 shows summary statistics of our outcome measures. The mean food safety expenditure per capita decreased from £3.13 in 2009/10 to £2.27 per capita by 2019/20. Over the same period, the average number FTE per 10,000 of the population in England reduced from 0.31 to 0.24 per 10,000 of the population. The average number of interventions achieved decreased by 13,246 per 100,000 establishments, and the average percent of broadly compliant establishments increased from 88% to 94 %.

**Table 2:** Summary table of Food Safety expenditure.

| Year | Average Food Safety Expenditure per capita (£) | Average number of Full Time Equivalent staff positions per 10,000 of the population | Average number of interventions achieved per 100,000 establishments | Average percent of broadly compliant establishments |  |  |
| --- | --- | --- | --- | --- | --- | --- |
|  |  |  |  | Average percent of broadly compliant establishments | Average number of broadly compliant establishments | Average number of establishments rated in FHRS scheme |
| 2009/10 | 3.14 | 0.31 | 73497 |  |  |  |
| 2010/11 | 3.20 | 0.30 | 71628 | - |  |  |
| 2011/12 | 2.85 | 0.29 | 69660 | - |  |  |
| 2012/13 | 2.79 | 0.28 | 68210 | 88% | 518 | 600 |
| 2013/14 | 2.66 | 0.27 | 66500 | 89% | 538 | 611 |
| 2014/15 | 2.45 | 0.27 | 64315 | 90% | 522 | 590 |
| 2015/16 | 2.44 | 0.26 | 64513 | 92% | 528 | 582 |
| 2016/17 | 2.34 | 0.25 | 62773 | 92% | 546 | 603 |
| 2017/18 | 2.32 | 0.25 | 62887 | 93% | 570 | 625 |
| 2018/19 | 2.30 | 0.25 | 62657 | 93% | 586 | 637 |
| 2019/20 | 2.27 | 0.24 | 60251 | 94% | 582 | 625 |

*Table 1 uses information adapted from Food Law Code of Practice, Annex 1 of Food Law Code of Practice, and the Food Standards Agency website (20,21)*

Figure 1 presents the percentage change in outcomes of interest relative to 2009/10. The number of FTE per 10,000 reduced by 23.5% in 2019/20. The number of interventions achieved per 100,000 establishments reduce by 18% by 2019. We see that from 2012 to 2018 the average number of broadly compliant establishments increased by 13 % before dropping to 12% in 2019/20 compared to 2009/10.

**Figure 1:**
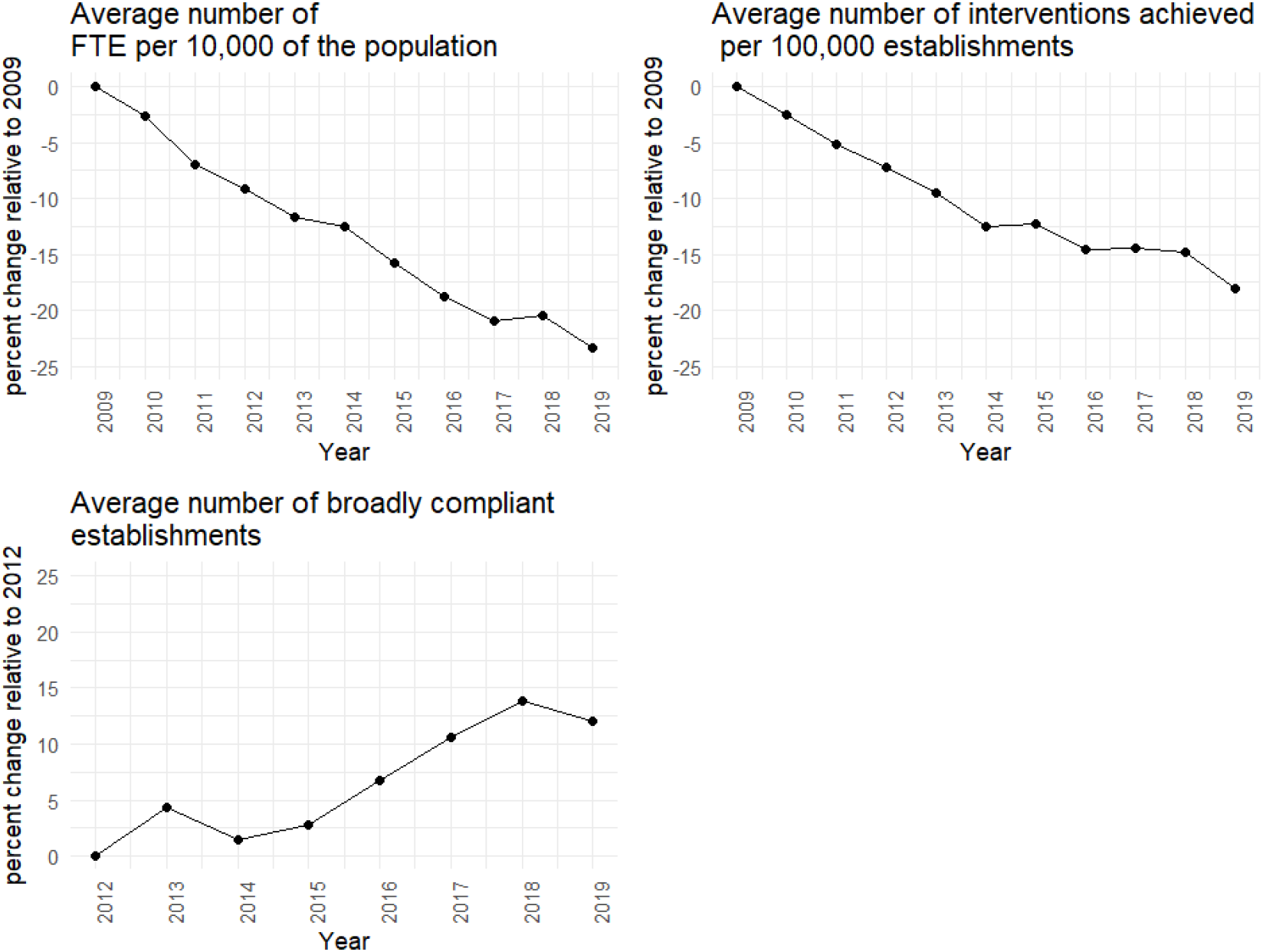
Percentage change in outcomes of interest relative to baseline year 2009 for measures of FTE, interventions and the proportion of compliant establishments. Year represents financial year

Table 3 presents the results from the regression analyses. The number of staff per capita decreased by 2.0% (p<0.01), with each £1 per capita reduction in food safety expenditure. The number of interventions per establishment saw a significant decrease of 1.7 % (p<0.05) with a £1 per capita decrease in food safety expenditure. For each FTE per 10,000 of the population decrease in staff, the number of interventions per establishment decreased by 49.2% (p<0.001). When accounting for number of FTE, a £1 reduction in expenditure was saw interventions decrease by 1.5% (p<0.05). When accounting for spend per capita, a reduction in staff was significantly associated with a decrease of 46.0% (p<0.001) in the number of interventions achieved per establishment. We identified no significant relationship between food safety expenditure or FTE and the odds of establishments achieving broadly compliant ratings. We carried out a sensitivity analysis on data with missing values present, the results of this can be found in supplementary material, section 4.

**Table 3:**
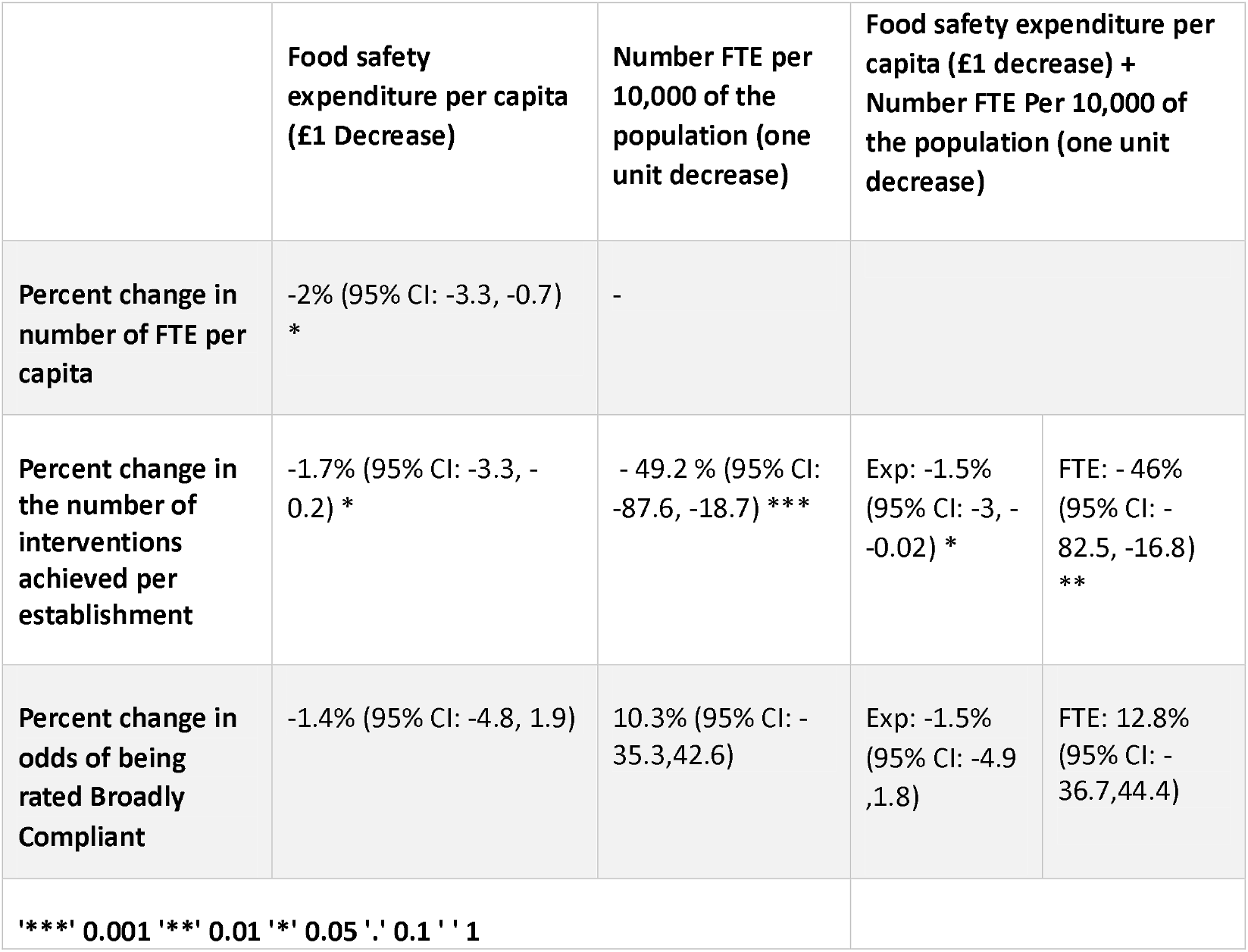
Regression analysis results showing effect of food safety expenditure and staffing levels on primary outcomes.

## Discussion

Our study shows for the first time how the reduction in local funding cuts had a substantial impact on the capacity of local statutory food safety services. Cuts had a negative impact on staff and the number of interventions achieved, but no significant relationship was found in relation to hygiene compliance.

Our findings are consistent with reports of staff cuts across public services in recent years due to austerity. Staff reduction is a key mechanism by which authorities try to make savings (12). This is reflected in a National Audit Office (NAO) report which reports food hygiene staff decreased by 13% between 2012/13 and 2017/18 (7), highlighting that even statutory services are vulnerable to cuts during austerity. This has been an approach taken by most authorities with 96% of single tier authorities and 86% of district councils reducing the number of staff between 2011 and 2013 (22). Some local authorities are reducing higher pay-grade staff, including managers (23). The loss of senior staff with more experience may create a workforce with less knowledge and expertise, a noted concern of some council workers (24). A survey by the Chartered Institute of Environmental Health described 56% of LA’s reported vacancies in EH teams as remaining unfilled for at least six months, primarily due to lack of qualified or experienced EHPs (25). Staff cuts and reduced ability to hire experienced staff into EHP roles may create a workforce which lacks depth and overall capacity to handle the rising demand of services.

Though important actions such as food hygiene inspections have been prioritized with spending cuts (26), our results show that a decrease in staff due to cuts are impacting the number of interventions achieved. The reduction in staff means the workforce is becoming more stretched with fewer staff having larger workloads (26,27) and with professionally qualified food hygiene staff carrying out more inspections and audits per person in 2019/20 compared to 2010/11 (9). In addition, local authorities failing to meet intervention targets noted staffing shortfalls as the reason and expressed concerns regarding the impact on food safety (7). They also noted changes in functions due to staff cuts, for example, cutting back on activities such as advice, guidance and sampling, prioritizing higher risk premises, and outsourcing to third party contractors to increase inspection capacity (7). This is in line with our results, and suggests a decrease in service capacity, which may increase the risk of foodborne disease outbreak.

Evidence shows that the capacity of EH teams is key for public protection, with workforce experience, workforce levels, and regulation compliance having protective effects against foodborne illness (28). Literature shows interventions such as training food handlers in food safety and hygiene are effective in reducing microbial prevalence in food service premises (29). Reducing access to advice and education visits, or other interventions, may see a decline in practice of food handlers and increase occurrence of microbial contamination, placing the public at greater risk of gastrointestinal infection. Previous work highlights the importance of EHP inspection visits, with 87.8% (n=387) of survey respondents in one study saying that the action taken following the inspection had improved the hygiene of their establishment (30). As a substantial amount of foodborne illness outbreaks are associated with settings that rely on intervention and guidance from EHP’s, our results are troubling. This is particularly true when considering the increasing trend of hospitalizations due to pathogens linked to foodborne disease (31,32)

While our results highlight challenges surrounding the capacity of food safety teams and service delivery, we saw no association between food safety expenditure or staffing levels and food hygiene compliance. Paradoxically, we see an increase in broadly compliant establishments during the study period, agreeing with other reports during this time (7). However, it is important to interpret these results with caution. While our results indicate that a reduction in resources does not impact hygiene compliance rates, there may be other factors involved. Such as the requirement of local authorities to achieve compliance as stated in the food law code of practice (21). Staff prioritize establishments of higher risk (26) meaning businesses achieving lower FHRS scores will likely be a resource priority to achieve compliance. The FHRS score offers a snapshot of the hygiene practice at time of inspection. While important, it provides limited detail concerning the hygiene practice of the establishments between inspections. There may be gaps in compliance by area or deprivation level, reflective of funding inequalities, exemplified in our analysis and previous research (10). Overall compliant food hygiene scores may show a level of resilience of food safety teams and FBO’s to resource cuts, however this does not eliminate the risk of outbreaks.

The reduction in expenditure and consequent trends take place in a time where demands are increasing as the number of registered food establishments are increasing (26). The existing inequalities in funding cuts to these services may lead to widening health inequalities as a result (10), with recent research showing reduced capacity in the more deprived areas for gastrointestinal infection work in the community (33). Evidence supports that investment in food and sanitation services is associated with the reduction in incidence of disease caused by key food and water related pathogens (34). Cutting of these services may be placing the public at greater risk to these infections.

## Strengths and Limitations

Unlike previous studies, we attempt to identify the extent to which reduced expenditure may impact services and their operation and capacity. We use data on key indicators of food safety in the context of food hygiene functions, to track trends. There are several limitations to our study. Firstly, we rely on ecological data, however this work allows us to explore important general trends of local funding cuts on these services across England. Secondly, this paper focuses primarily on food safety services. Whilst this is important, there are also other routes they GI infections can spread in which these services will not influence. Such as person-to-person transmission within communities, transmission from contact with animals, or exposure whilst travelling outside of England. Thirdly, there were limitations to data quality, such as missing data. To mitigate this issue, we used multiple imputation. Additionally, reduced expenditure and staffing could have impacted data collection and quality, such as with compliance, potentially introducing bias. The funding cuts may have altered data collection and may, in part reflect altered operational practices in time of fiscal constraint. Finally, directly relating expenditure decrease to FTE outcomes may oversimplify results, as overhead allocation and fixed departmental costs were not accounted for.

## Conclusion

In this study we quantify the impact of food safety expenditure reductions on indicators of food hygiene functions, crucial for prevention of foodborne infectious disease. Whilst ER and food safety and infection control services have seen changes in service expenditure overtime, there is little to no knowledge of the association between local funding cuts and impact and service delivery. We show here that decreases in food safety expenditure significantly impacted staffing levels and service provision. Whilst this raises concern about the capacity of food safety teams to protect the public, we find no impact on hygiene compliance. Our work raises questions about the impact these trends on public risk to gastrointestinal infection, and any inequalities that may be present.

## Data Availability

All data produced in this study are available upon request

https://pldr.org/dataset/2omjn/cultural-environmental-regulatory-and-planning-services-individual-spending-lines-r05-fin0761

https://www.nationalarchives.gov.uk/webarchive/

## Acknowledgements

We gratefully acknowledge the contributions of Alexandros Alexiou, David Hughes, Simon Melican, Samantha Walters, Jane Muizelaar,

## Contributors

LM, HEC, RG, XZ, MAG, MAC, IEB, BB and DH were involved in conceptualisation of the study. LM and DH were involved in investigation, and project administration. LM, DH, HEC, MAG, BB, IB and XZ contributed to methodology. LM carried out visualisation, writing–original draft preparation. LM, HEC, RG, XZ, MAG, MAC, IEB, BB and DH contributed to writing–reviewing and editing. HEC, XZ, MAG, MAC, IEB, BB and DH contributed to supervision. BB was responsible for funding acquisition. LM accepted full responsibility for the finished work and/or the conduct of the study, had access to the data and controlled the decision to publish. LM is responsible for the overall content as guarantor.

DH is currently in receipt of research grant support from the Food Standards Agency. IB was AstraZeneca’s Chief Data Scientist Advisor 2019-2023. LM, RG, MAC, HEC, XZ, MAG, and BB declare no relevant competing interests.

Ethics approval is not applicable, data used in this study is open access available to the public or available upon freedom of information request.

