## Supplementary material for "Understanding the impacts of reductions in local government expenditure on food safety services, England 2009/10 - 2019/20": Online Supplemental material

### 1.Data

#### Table S1.1 Data sources used in the paper

| Variable | Source |
| --- | --- |
| FTE(LAEMS) | Open access: <https://www.data.gov.uk/dataset/090b5b23-5020-4480-96a0-8b294ca82653/local-authority-food-law-enforcement-returns> |
| Number of interventions achieved (LAEMS) | FOI request from FSA |
| FHRS score (FHRS) | Open access at time of access, now access request needed CDRC |
| Food Safety expenditure | Open access: <https://pldr.org/dataset/2omjn/cultural-environmental-regulatory-and-planning-services-individual-spending-lines-r05-fin0761> |

#### 1.2 Food hygiene interventions

Local authority service indicators for food safety were sourced from FSA’s local authority enforcement monitoring system (LAEMS). This is a system in which local authorities report data from food law enforcement activities (1) for food standards and food hygiene. Food standards refer to activity in relation to the labelling, presentation, composition, and overall quality of the food (2). Food Hygiene data relates to regulatory activity of microbial contamination of food (3) and foodborne illness. The FSA collects data on these activities on an annual basis to monitor food law enforcement (4). In this study we analyse annual data on food hygiene returns only. The reporting year runs from 1st April to 31^st^ March the following year.

Measures derived from the Local Authority Enforcement Monitoring System (LAEMS) include the number of interventions achieved per local authority each year. Interventions are carried out routinely and are scheduled using a risk-based approach. Risk ratings are given following an initial inspection (5), with the risk rating guiding the type of intervention that can be carried out. The risk rating represents the risk faced to the public and is derived from the type of food and method of handling, method of processing, consumers at risk, level of current compliance, vulnerable groups and risk of contamination (6). Higher risk establishments will receive interventions more frequently. These will include inspections and audits. Lower risk establishments will receive interventions less frequently and may receive alternative interventions or inspections and audits.

##### *1.3 Local authority enforcement monitoring scheme FTE data*

It is advised that the number of FTE reflect the proportion of time spent by staff on food hygiene issues (7). However, there is no prescriptive guidance on how the time spent on enforcement should be determined, therefore figures supplied are estimates (7). Due to the variety of methods on calculating time the FSA only use the data to compare year on year to look at overall trends in FTE across the UK (7). Here in this paper, we do not make direct comparisons between LA’s but analyse general trends across time, complying with guidance by FSA.

Calculation of FTE: The number of FTE represent the number of professional or administrative staff posts allocated to food law enforcement work and are separated by responsibility e.g. food hygiene or standards (6). LAEMS provide an example of how the FTE are calculated, this is included below:

“*five professional posts were allocated to carry out food hygiene work during the year, and three of these posts were filled for the full twelve months, one for six months and a contractor was employed for 3 months, the figures reported should be as follows: FTE posts allocated = 5; FTE posts occupied = 3.75 i.e. 3 posts filled for full twelve months + one filled for six months (0.5) + one contractor employed for three months (0.25). Where a professional and/or administration staff member only spends a proportion of their time on food hygiene and/or food standards issues, the calculation should reflect this. We recognise that the figures supplied will often be ‘educated estimates’.” (Local authority enforcement monitoring system 2019*(6)

#### 1.4 Food Hygiene Rating Score ratings 1-5

Establishments can receive any of the below scores after inspection.

0. Urgent improvement is required
1. Major improvement necessary
2. Some improvement necessary
3. Hygiene standards are generally satisfactory
4. Hygiene standards are good
5. Hygiene standards are very good (8)

Extra information on FHRS scheme <https://www.food.gov.uk/business-guidance/how-food-hygiene-ratings-work>

### 2 Missing data and data excluded

#### Table S2.1Local authorities excluded from analysis

|  | **LA’s removed due to >50% Missing** | **LA’s removed due to reporting with food standards** | **LA’s removed due to inconsistent reporting** |
| --- | --- | --- | --- |
| 1 | Stratford Upon Avon | Westminster | Adur |
| 2 | Rutland | Hillingdon | Worthing |
| 3 | Rochdale | Bexley | Rother |
| 4 | Medway | Hartlepool | Wealden |
| 5 | Tameside | Bolton | Babergh |
| 6 | High peak |  | Midsuffolk |
| 7 | Herefordshire |  |  |
| 8 | Hambleton |  |  |
| 9 | Craven |  |  |
| 10 | Portsmouth |  |  |
| 11 | Epsom and Ewell |  |  |
| 12 | Hinckley Bosworth |  |  |
| 13 | Hart |  |  |

*Note: The following local authorities provided no service expenditure data for the specified years; Havant 2017, East Hampshire 2017, Newark and Sherwood 2012, 2013 and Folkestone Hythe for 2017 and 2018.*

*Adur, Worthing, Babergh. Midsuffolk, Rother and Wealden were also excluded due to inconstant reporting level*

It was identified that negative expenditure was present in a small number of local authorities. We treated these data as 0/missing after contacting a sample of the authorities and identifying mixed reasons as to why there may be negative values reported including error, and system calculations and general uncertainty. Therefore, we felt the best course of action would be to treat the data as missing for multiple imputation models.

The total number of local authorities with missing data (0 or NA) for the food safety expenditure, FTE, number of interventions, number of establishments/ businesses rated for FHRS were tallied.

### 3. Trends in Food safety expenditure by deprivation, local authority structure and rural or urban categorisation

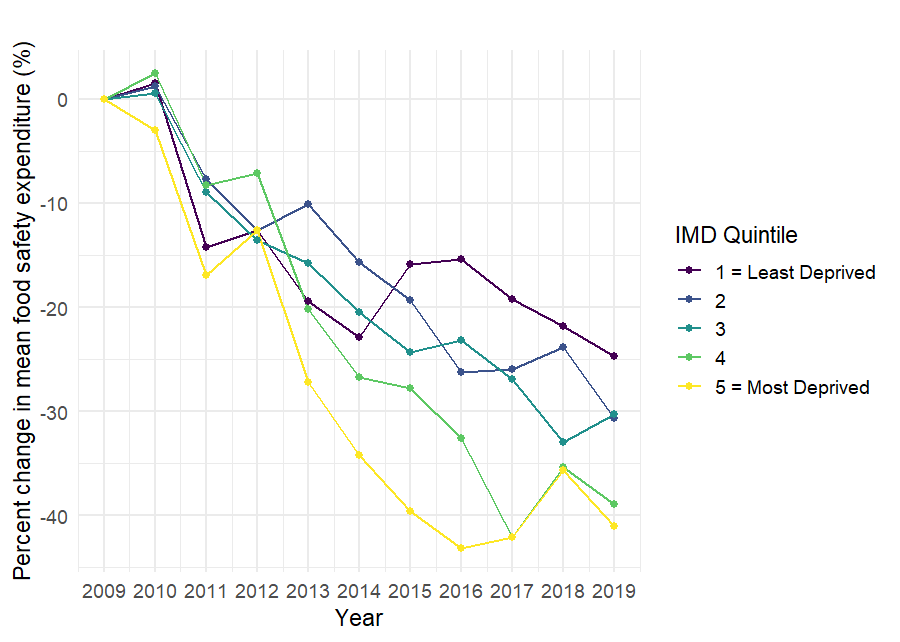

*Figure S3.1 shows the percentage change in food safety expenditure per capita relative to 2009 by quintiles of deprivation*

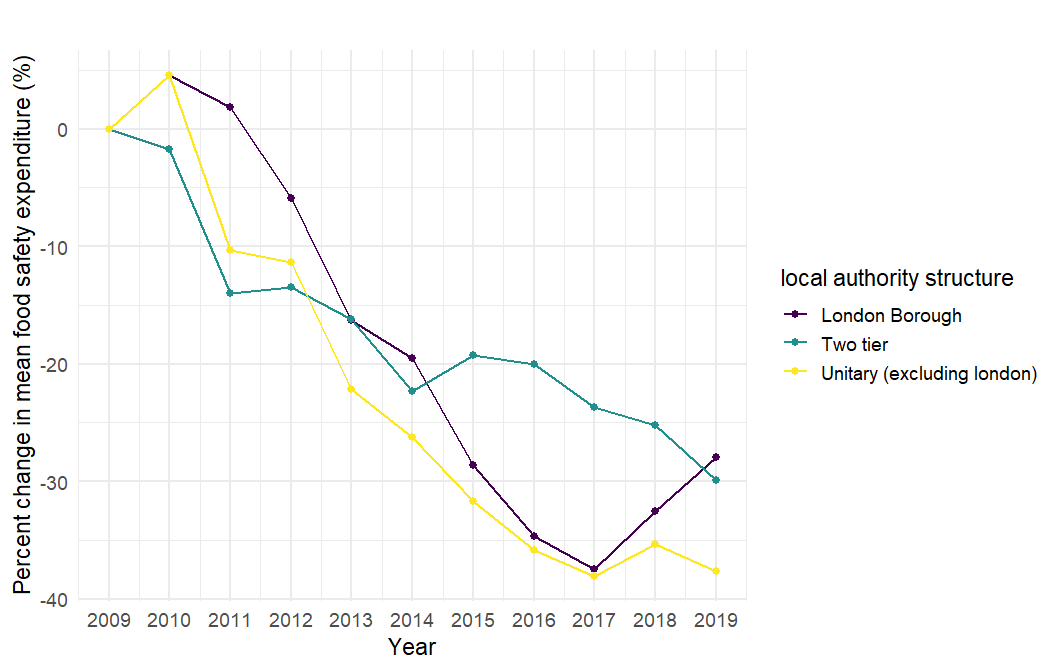

*Figure S3.2 shows the percentage change in food safety expenditure per capita relative to 2009 by local authority structure*

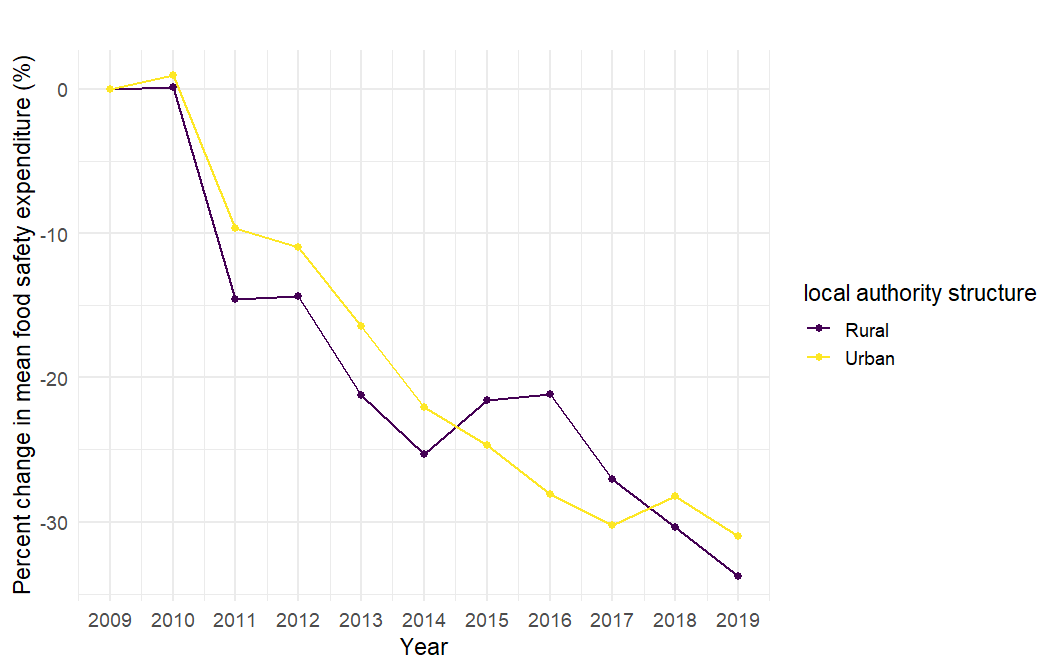

*Figure S3.3 shows the percentage change in food safety expenditure per capita relative to 2009 by rural or urban*

### 4. Sensitivity analysis

We carried out a sensitivity analysis, this data covers 312 authorities and includes local authorities with NA present.

#### Table S4.1 Sensitivity analysis regression models

|  | Food Safety Expenditure per capita (£1 Decrease) | Number FTE Per 10,000 of the population (One Unit Decrease) | Food Safety Expenditure per capita (£1 Decrease) + Number FTE Per 10,000 of the population (One Unit Decrease) | |
| --- | --- | --- | --- | --- |
| Number FTE Per 10,000 of the population (One Unit Decrease) | -2.4 % (-3.8, -1) *** | - |  | |
| Number of Interventions Achieved | -1.9% (-3.2, -0.6) ** | - 51.4%) -89.3, -21.1) *** | Exp: -1.5% ( -2.8, -0.3) * | FTE: - 39% (-72, -13) ** |
| Proportion of Broadly Compliant Establishments | -1.2% (-4.1,1.6) | -7.9 (-39.4, 39.9) | Exp: -1.2% (-4.1, 1.8) | FTE: 7.8% (-40.5,39.4) |
| '***' 0.001 '**' 0.01 '*' 0.05 '.' 0.1 ' ' 1 | | |  | |

5.	Food law code of practice (England). :101.

6.	laems guidance [Internet]. Available from: chrome-extension://efaidnbmnnnibpcajpcglclefindmkaj/https://old.food.gov.uk/sites/default/files/laemsguidance_0.pdf

7.	Local Authority Food Law Enforcement returns - data.gov.uk [Internet]. [cited 2024 Dec 16]. Available from: https://www.data.gov.uk/dataset/090b5b23-5020-4480-96a0-8b294ca82653/local-authority-food-law-enforcement-returns

8.	Food Hygiene Rating Scheme | Food Standards Agency [Internet]. [cited 2024 Mar 16]. Available from: https://www.food.gov.uk/safety-hygiene/food-hygiene-rating-scheme
